# Transmission routes of Covid-19 virus in the Diamond Princess Cruise ship

**DOI:** 10.1101/2020.04.09.20059113

**Authors:** Pengcheng Xu, Hua Qian, Te Miao, Hui-Ling Yen, Hongwei Tan, Min Kang, Benjamin J. Cowling, Yuguo Li

## Abstract

**Background:** An outbreak of COVID-19 occurred on the Diamond Princess cruise ship in January and February 2020. We analysed information about cases to infer transmission dynamics and potential modes of transmission.

**Methods:** We collected the daily number of 197 symptomatic cases, and that of the 146 passenger cases in two categories, i.e. those who stayed and did not stay in the same stateroom. We retrieved the quarantine details and the ship’s 14-day itinerary. We searched the websites of national/local health authority along the cruise routes and local news using Google for locally confirmed cases associated with the ship. We obtained the design of air conditioning and sewage treatment of the ship from literature. We back-calculated the dates of infection from the epidemic curve and compared with the start of on-board quarantine.

**Results:** Major infections started on Jan 28 and completed by Feb 6 for passengers except those who stayed in the same stateroom with infected individual(s). No other confirmed cases were identified among the disembarked people in Hong Kong except an 80 years old passenger. No confirmed cases were reported in three other stopovers between Jan 27-31 associated with disembarked passengers or visitors from the ship, however two Okinawa taxi drivers became confirmed cases in association with driving the ship passengers. Infection among passengers after Feb 6 was limited to those who stayed in the same stateroom with an infected passenger. Infections in crew members peaked on Feb 7, suggesting significant transmission among crew members after quarantine on Feb 5.

**Conclusions:** We infer that the ship central air conditioning system did not play a role, i.e. the long-range airborne route was absent in the outbreak. Most transmission appears to have occurred through close contact and fomites.

**Significance Statement:** Transmission by the long-range airborne route for SARS-CoV-2 in the 2020 Diamond Princess Covid-19 outbreak has been debated with significant implication for intervention. We found that the transmission by close contact and fomite explains the outbreak, and the central air-conditioning system did not play a role, demonstrating the importance of social distancing, good hygiene and maintaining good building ventilation for intervention.

## Introduction

Uncertainty remains on the transmission routes of SARS-CoV-2 as the current Covid-19 pandemic virus continues to spread all over the world, cause mortality, and significantly disrupt human life and economy.

The large Diamond Princess Cruise outbreak in Japan as impacted by quarantine may shed light on the possible transmission routes, and the roles played by close contact, fomites and central air-conditioning system of the ship.

By Mar 5 2020, a total of 696 cases (Sanki, 2020) were confirmed among the passengers and crew in the Diamond Princess Cruise ship, which had 3711 persons (2666 passengers and 1045 crew members) (Nishiura, 2020). Among the confirmed cases, 552 were passengers, 144 crew members, and 410 had no symptoms. On Jan 20, an 80-years old man X become a passenger of the ship in Yokohama. He had fever on Jan 23, disembarked at the port of Hong Kong on Jan 25 and confirmed to be infected with Covid-19 on Feb 1. The ship continued her voyage by visiting four ports in the region and returned to Yokohama, Tokyo, Japan on Feb 3 earlier than its scheduled date of Feb 4. The ship was quarantined by the Japanese Government at sea, beginning at 7 am, Feb 5 with 2,666 passengers being asked to stay in their stateroom and 1,045 crew on board. The quarantine ended on Feb 19. On Mar 1, all passengers and crew were disembarked.

In general, a cruise ship in a voyage is like a small town or community. As the passengers and crew sleep, eat and play, a cruise ship is probably one of the most crowded 24 hr human-made public communities. Large outbreaks of norovirus were frequently reported in cruise ships (CDC, 2020a). Cruise ships may be considered as a place integrating all possible crowded indoor environments on land – restaurant, swimming pool, casino, theatre, bars, food streets, hotels, etc. For the Diamond Princess Cruise ship during its voyage, all these on-board public spaces were open to all passengers on Jan 20-Feb 4, but suddenly closed when the Japanese Government implemented the 14-day quarantine from 7 am, Feb 5. During the quarantine period, passengers stayed in their rooms, and the outer cabins had access to balcony doors. The transmission routes of the virus may be pinpointed by examining who were infected before and during the quarantine period. For example, as passengers stayed in their individual staterooms, transmission by the recirculating central air conditioning systems may be implied if there were cross-room transmission between passengers.

## Methods

### Data

We collected the daily number of 197 symptomatic cases from Jan 20 to Feb 18 and retrieved data of the implemented quarantine measures and implementation schedules from the Ministry of Health, Labour and Welfare, japan website (MHLW, 2020a). We also extracted from the same website the data of 146 passenger cases including 129 in the “close contact” category, i.e. those who stayed in the same stateroom with infected individuals (s) and 17 in “non-close contact”, i.e. those who did not stay in the same stateroom with infected. We obtained the itinerary of the ship from Yutaka Club Cruises International (2020). We searched the websites of national/local health authority in Vietnam, Hong Kong and Taiwan, and their local news using Google along the cruise routes for locally confirmed cases associated with the ship. We also obtained the basic data of the ship such as space ratio, number and size of staterooms from Cruisedeckplans (2020). The air conditioning design of the ship was obtained from Kosako and Shiiyama (2008) and the waste and sewage treatment system design from Onoguchi et al (2004).

### Estimation of infection time

We estimated the daily number of exposed (infected) individuals who had received a sufficient dose of the SARS-CoV-2 virus to develop disease symptoms (defined as ‘symptomatic cases’). Let *E*_*n*_ be the number of exposed (infected) cases on day *n, S*_*n*_ the daily number of individuals with onset of symptoms and *ρ*_*j*_ the probability of the onset of symptoms on the *j*th day after exposure, where *ρ*_*j*_ follows a log-normal distribution (Li et al., 2020).

The daily numbers of infected cases and new symptomatic cases are related by the following equation:

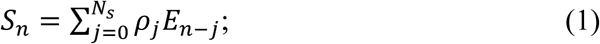

where *N*_*s*_ is the longest incubation period from infection (exposure) to symptom onset. Rewriting this equation in a matrix form for the entire study period of *N* days, we have Equation (2):

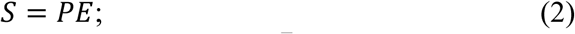

where *S* = [*S*_1_, *S*_2_ … *S*_*N*_]^*T*^ and 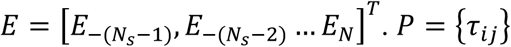 is an *N* × (*N* + *N*_*s*_) matrix with 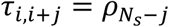 for 1 ≤ *i* ≤ *N*, 1 ≤ *j* ≤ *N*_*s*_; otherwise *τ*_*i,i+j*_ = 0.

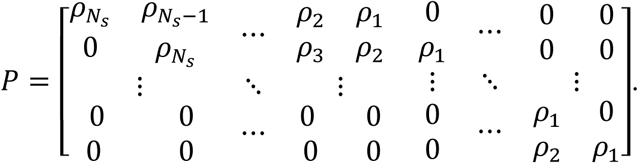

Equation (2) is ill-posed. The daily number of infected cases, *E*_*n*_, can be obtained by ensuring the following constraints.

- Non-negative condition, *E*_*n*_ ≥ 0, as the daily number of infected cases cannot be a negative number.
- Conservation condition, 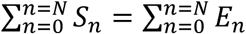: the total number of infected n=0 n=0 individuals must be equal to the total number of individuals displaying symptoms.
- Modulus minimum condition (Tikhonov regularisation): we choose the solution that minimises the modulus, min ‖*E* ‖^2^.

Following these constraint conditions, we define the extended symptom-onset matrix S̃ = 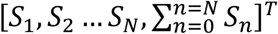; and the extended incubation period matrix as

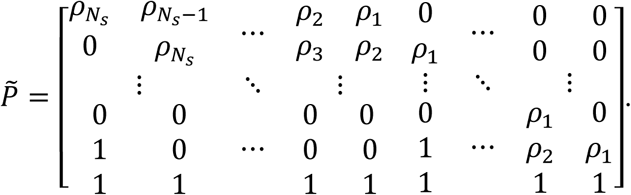

We then obtain the following equation

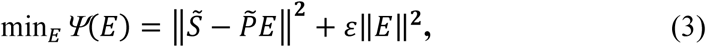

where *E*_*n*_ ≥ 0.

The choice of parameter *ε* affects the numerical solutions; parametric analyses (Supplementary Information) indicated that the use of *ε* = 0.005 afforded the best solutions.

The steepest descent method was used to solve the above nonlinear problem. An early version of this approach was combined with an approximation method to estimate the infection rate of general infection spreaders during the 2003 SARS epidemic. The method predicted the reported occurrence of all seven SARS super spreading events in Hong Kong and Singapore in 2003 (Li, et al., 2004), in terms of both exposure times and number of infected cases. The validation of the method for Covid-19 is shown in Figure S1.

## Results

Figure 1a shows the predicted daily infection for both the 149 passengers and 48 crew. The infection probably started on Jan 28 and completed by Feb 6 for passengers (Figure 1a). However, crew members continued to get infected on and after Feb 6. The infection started among the passengers with a peak on Feb 3, which was the date when the ship just arrived in Yokohama. The decline of the infection after Feb 3 is expected as some passengers probably started to minimize their close contacts and social gathering. The crew infection was peaked on Feb 7, suggesting that some close contact continued to be significant among crew members after quarantine on Feb 5. This is not surprising as the crew members need to provide serves to the passengers. Kakimoto et al (2020) described the transmission among crew members during quarantine of the ship. The 3-4 day late peak infection date for the crew also suggest that infection was initiated first among the passengers, as also suggested by the study of Kakimoto et al (2020).

**Figure 1.**
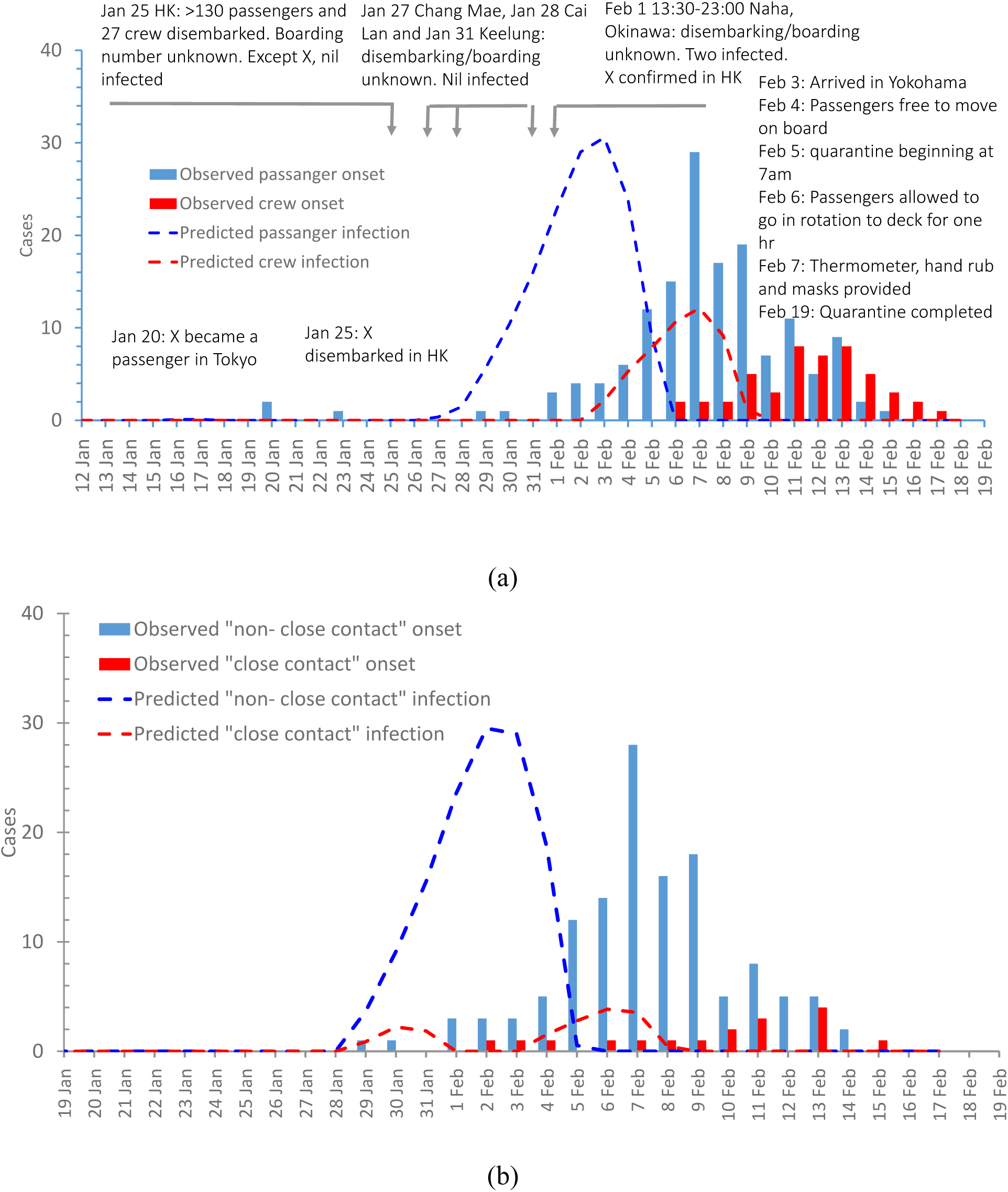
(a) Predicted infection dates and observed symptom onset dates for 149 passengers and 48 crew; and (b) for those infected due to close-contact (staying the same stateroom with another infected) and non-close-contact (not in the same stateroom). Note that some close contact passenger cases were also infected between Feb 6-7, differ from all passenger prediction in (a) in which no infection on Feb 6-7 were predicted.

Categorizing the 146 infected passengers into “close contact” and “non-close contact”, we find that those “close contact” infection occurred after the quarantine starting date of Feb 5. Note that the original figure caption of (https://www.mhlw.go.jp/content/10200000/Fig2.pdf) stated the data was for 149 passengers, but the data shown in the figure was actually for 146. These “close contact” infection was among those who stayed in the same stateroom with the infected individuals (Figure 1b). There seems to be two waves of infection within the “close contact” passengers. The number of “close contact” infected passengers was small as each stateroom mostly had a maximum of 4 persons (Cruisedeckplans, 2020).

The chronological events associated with the outbreak are summarized in Figure 2. We first found that there was no secondary infection reported at all ports after the ship stopped over in its 14-day round-trip voyage prior to arrival in Yokohama except in Okinawa. More than 130 passengers and 27 crew members disembarked in Hong Kong on Jan 25 together with an 80 years old X (Case no 14 in Hong Kong, CHP, 2020). No other confirmed cases were identified among these disembarked passengers (86 Hong Kong residents) and crew members in Hong Kong, suggesting that transmission in the cruise ship during Jan 20-25 was limited, which agrees with the observed onset data and the predicted infection data (Figure 1A). The ship stopped over in Chang Mae and Cai Lan on Jan 27 and Jan 28 respectively, and in Keelung on Jan 31, no confirmed cases were reported in all three locations associated with disembarked passengers from this ship.

**Figure 2.**
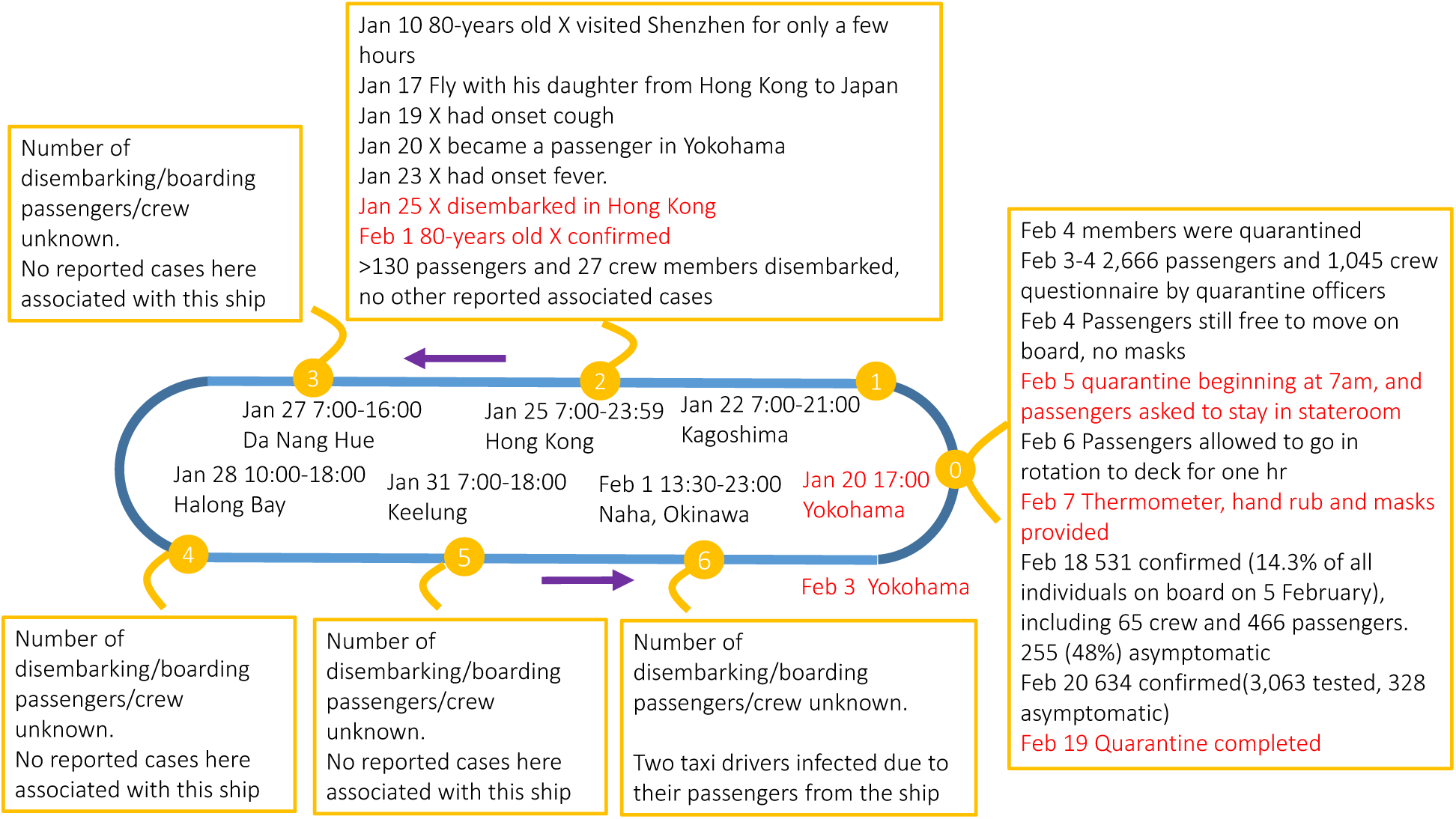
Chronological events associated with the Diamond Princess Covid-19 outbreak on Jan 22-Feb 20. Compiled using information from websites of local/national health authorities from Japan, Hong Kong, Vietnam and Taiwan.

The significantly low number of infection among passengers after Feb 6 is in line with the imposed quarantine starting at 7 am, Feb 5 such as the stay-in-room policy, and the availability of masks on ship from Feb 7; see the left hand side box in Figure 2. Infection among passengers after Feb 6 was then limited to “close contact”, i.e. those who stayed in the same stateroom with infected (Figured 1b).

## Discussion

The difference in our predicted infection prior to and after Feb 5 quarantine in the Diamond Princess Cruise Ship outbreak reveal the possible transmission routes involved in. For those infected in staterooms without an infected individual (i.e. non-close contact category), the predicted infection all occurred prior to Feb 5. The peak effective reproduction number on the ship was approximately 11 as predicted by Mizumoto and Chowell (2020), much higher than the mean estimates elsewhere of 1.7-7 (Mizumoto and Chowell, 2020). However, they also predicted a peak infection 1-2 days after Feb 5, which differs slightly from our predicted peak on Feb 3. The infection among passengers was significantly reduced following the quarantine starting on Feb 5, but according to our prediction, the passenger infection was already on the way to decline from 3 Feb as they became aware of the outbreak and exercised self-discipline. For those infected in the staterooms with infected individual (i.e. close contact category), the infection occurred both prior to and after Feb 5 quarantine. The lack of cross-room transmission between passengers of different staterooms during the quarantine period strongly suggests that the ship central air conditioning system did not play a role, i.e. the long-range airborne route was almost totally absent in the outbreak, based on our predicted data. The Diamond Princess Cruise Ship was built and completed in 2004 by Mitsubishi Industries. A centralized full-air air conditioning system with VAV was designed and installed in all passenger staterooms and crew cabins, with 30% fresh outdoor air for staterooms, 50% for public areas and 100% for clinics and kitchen etc. A fully independent exhaust air system was installed from the bathrooms. Although no measured ventilation rate was reported for the ship, international standard such as ISO 7547:2004 requires 8 Litre/s per person, which is similar to those in offices and other public spaces (ASHRAE62.1, 2007). It was also reported that the maximum outdoor air supply was operated during the quarantine period. This probably implied that a reasonably ventilated indoor environment such as that on this ship would not result in cross transmission of the virus due to the long-range airborne route.

On the other hand, the predicted higher infection of non-close contact passengers prior to the quarantine and the higher infection for crew after the quarantine strongly suggested that the close contact and/or fomite transmission played a role. Figure 1b shows that the non-close contact transmission of beyond-room passengers all occurred prior to Feb 5. The number of the infected in the close contact category, i.e. within-cabin transmission is only 17, the predicted infection dates might not be sufficiently accurate, and the data presented in Figure 1B is only indicative. The peak infection date of the within-cabin or close contact infected is clearly behind that of the non-close or beyond-cabin contacted.

The infection settings prior to the quarantine on Feb 5 might be similar to those norovirus outbreak reported in the cruise ships. Diamond Princess actually had an outbreak of gastroenteritis caused by norovirus in Feb 2016, in which 158 passengers and crew were sick on board (ABC News, 2016). It is known that the norovirus infections on cruise ships affected the largest number of people (Harris et al., 2010), often at hundreds. Crowdedness has been cited as an explanation for this cruise ship norovirus infection phenomenon (Harris et al., 2010, and Bert et al., 2014). Surfaces on cruise ship public restrooms are known to have high potential for fecal contamination (Carling et al., 2009). The same effect of crowdedness and fomite for the norovirus might have also happened to SARS-CoV-2 in this ship. The crowdedness and close contacts due to on-board leisure and social activities outside the staterooms could explain the observed higher infection among non-close contact passengers prior to the quarantine. According to the National Institute of Infectious Diseases (NIID), Japan website (NIDD, 2020), for the 536 Covid-19 cases among 2646 passengers, a higher cabin occupancy corresponds to a slightly higher attack rate, possibly due to the relatively smaller number of the within-cabin close contact transmission. Based on our retried data from a drawing, only 6 out of 75 passengers from 1-person cabins became confirmed cases (attack rate 8%), while for 2-person cabins, 485 of 2386 passengers were infected (20%), for 3-person cabins, 27 out of 125 (22%) and for 4-person cabins, 18 out of 60 (30%). The spatial distribution of the infected cabins can also reveal if the central air conditioning played role. Among the 1353 staterooms available on board, the cabin area ranges from 15-16 m^2^ for 1137 “interior”, “oceanview” and “balcony” rooms, 26 m^2^ for 186 “mini suite” rooms, 31 m^2^ for 29 suites or family suites, and one grand room of 58 m^2^. We do not have the data of distribution of the confirmed cases among these rooms. Sekizuka et al (2020) stated that “all COVID-19 patients were widely distributed in all 18 decks across the ship”. The spatial distribution of the confirmed cases in another cruise ship outbreak involving a small number of the infected passengers might also serve as a reference (International Financial News and Beijing News, 2020). The World Dream ship departed from Port of Nanshan, Guangzhou on 19 Jan 2020, visited Nha Trang and Da Nang in Vietnam on 19-24 Jan with 6903 passengers and crew on board, and among them, 108 were from Hubei in which 28 from Wuhan. The ship returned to the port of Nansha on 24 Jan. The distribution of the nine infected staterooms and the distribution of the 183 Hubei passengers on the ship were both random. Central air-conditioning system is commonly designed in spatial zones, and each air handling unit serve one zone. We failed to identify any spatial infection clusters in the World Dream cruise ship outbreak.

Our study thus offers some support for the current recommendation by most health authorities that the COVID-19 virus is mainly transmitted by close contact and fomite (WHO, China NHC, US CDC, 2020).

We reveal that most infection in the ship occurred during a period when passengers and crew members were still in regular or often close contacts before the quarantine began. The on-board transmission must have been rather limited prior to Jan 28. This explained why none of the more than 130 passengers and 27 crew members who disembarked in Hong Kong on Jan 25 developed the illness, and also no on-land infection was reported on Chang Mae and Cai Lan in Vietnam, and also at Keelung. 77 (59.7%) of the 129 infections among the passengers occurred during the ship’s journey between Okinawa and Yokohama, i.e. on Feb 2-4. It is noted that significant infection only started to grow on board from Jan 28 (Figure 1a). The on-board number of the symptomatic cases on Jan 31 was not significant. This probably explains the lack of infection of any local residents at the port of Keelung. Interestingly, two taxi drivers were confirmed in Okinawa respectively on Feb 14 and Feb 19 (MHLW, 2020b) and both had the ship passengers as their customers on Feb 1. The possible infection date of Feb 1 of the two taxi drivers also agrees with the significant number of infected individuals on the ship, though majority were pre-symptomatic or asymptomatic then. We do not have access to the passenger/crew boarding and disembarking data at the other 4 ports except Hong Kong for further analysis.

If there was a lesson to be learned, the ship could immediately disembark all passengers and quarantine them on land on Feb 1 when the former 80 years old passenger confirmed his infection, which might be considered as too idealistic. Our prediction shows that the implemented quarantine at 7 am Feb 5 worked well for the passengers, but not for the crew probably due to the need of passenger service by the crew (refs). There is a need for improved prevention measures for crew in similar situations.

Our estimation method is not without limitations. We cannot rule out the possibility of other infections during later part of the quarantine period due to incomplete data. Among the 634 cases confirmed by Feb 20, though 328 were asymptomatic, 306 were symptomatic at the time of confirmation. However, only 197 of the 306 symptomatic individuals had onset date, and further data from the remaining 109 symptomatic individuals may change our prediction.

Our predicted data also did not include possible transmission between crew and passengers prior to and during the quarantine period as they come into contact with each other, such as serving meals. We used the estimated incubation period from 186 patients in China, with known dates of both infection and onset of symptoms. The governing equation is known to be ill-posed, and assumptions were made (see Methods section, and also Li et al, 2005), though our method successfully predicted all seven reported super spreading events in the 2003 SARS epidemics in Hong Kong and Singapore. Figure S1 also showed that we also successfully predicted in the infection date for the 186 patients. Lastly, our approach cannot be used to study the roles of the asymptomatic cases in the transmission. The asymptomatic cases were first detected on the ship when the tests were extended to all passengers on Feb 15 after focusing in those with symptoms (Wikipedia, 2020). If the spatial distribution (room where the confirmed case stayed) of both symptomatic and asymptomatic cases is available, it may be possible to infer if any asymptomatic transmission occurred and how.

## Data Availability

Not available right now

## Acknowledgements

This work was supported by the Research Grants Council of Hong Kong’s General Research Fund (grant number 17202719) and Collaborative Research Fund (grant number C7025-16G).

## Author contributions

Y.L. and H.Q. contributed to the conception and design of the study; P.X. contributed to the modelling, Y.L., H.Q., T.M., M.K. and H.T. contributed to the data collection and analyses, H.-L.Y., B.J.C. M.K and H.T. contributed to analyses and interpretation. Y.L. wrote the manuscript and all authors contributed to the revision.

All authors approved the submitted version and have agreed to be personally accountable for the authors’ own contributions.

## Supplementary Appendix

### Validation of our back calculation method

We extract 186 confirmed cases with definite exposure date and symptom onset date, from a dataset collected from individual municipal health authorities in non-Hubei provinces in the Mainland China. These 186 cases were used to validate our prediction method, first to obtain an incubation period distribution, followed by comparison of the predicted onset from the infection data, and comparison of the predicted infection from the onset data.

**Figure S1.**
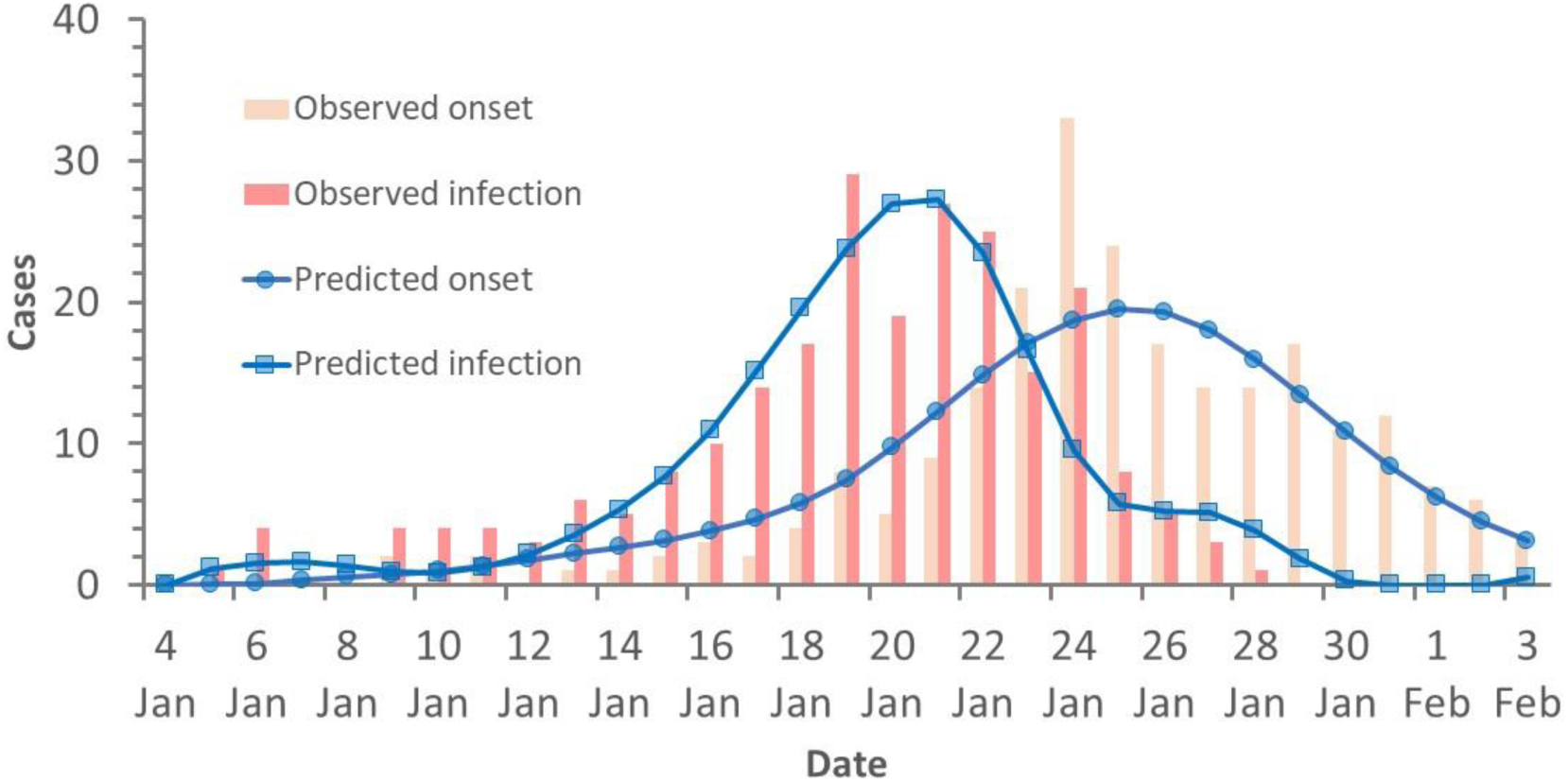
Comparison of the predicted onset from the infection data, and comparison of the predicted infection from the onset data for the 186 confirmed cases with definite exposure date and symptom onset date, from a dataset collected from individual municipal health authorities in non-Hubei provinces in the Mainland China.

### Comparison of available data in the literature

We found three sets of onset data of passengers for the Diamond Princess outbreak in two categories, i.e. close contact and non-close contact.

**Table S1.**
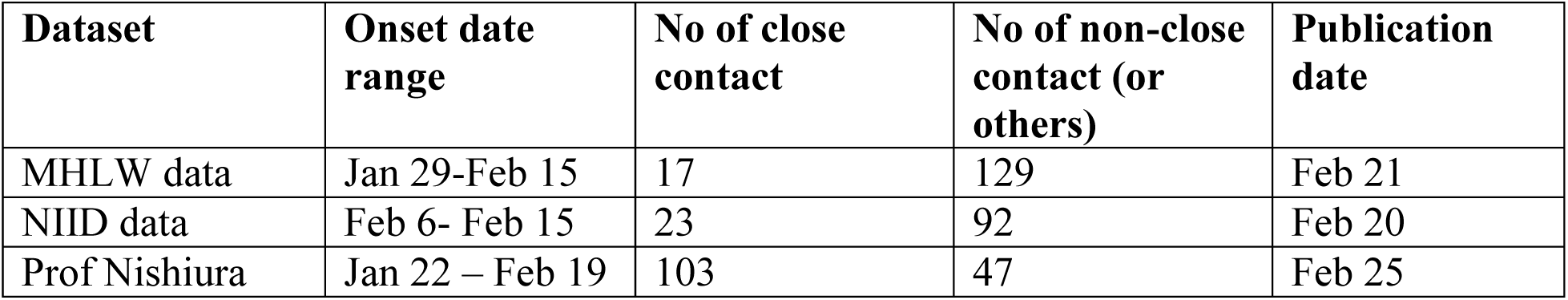
A summary of the three data sets of the passenger onset data in the Diamond Princess outbreak

| Dataset | Onset date range | No of close contact | No of non-close contact (or others) | Publication date |
| --- | --- | --- | --- | --- |
| MHLW data | Jan 29-Feb 15 | 17 | 129 | Feb 21 |
| NIID data | Feb 6- Feb 15 | 23 | 92 | Feb 20 |
| Prof Nishiura | Jan 22 – Feb 19 | 103 | 47 | Feb 25 |

#### Onset data 1

MHLW data: This was presented in a Press Conference on the Situation of the novel Coronavirus Disease in the Cruise Ship off the coast of Yokohama, held in the MOFA Press Conference Room from 20:40 to 21:40 on Friday 21 February 2020. Prof NISHIURA Hiroshi was one of the presenters.

Figure 2 in the original presentaton: Number of COVID-19 cases with symptoms among the passengers on the Cruise ship (by close contact status, as of 02/19/2020) https://www.mhlw.go.jp/content/10200000/Fig2.pdf

##### Note from the authors of this paper

There were 146 cases though on the figure legend stated that there were 149 cases.

#### Onset data 2

NIID data: Field Briefing: Diamond Princess COVID-19 Cases, 20 Feb Update.

https://www.niid.go.jp/niid/en/2019-ncov-e/9417-covid-dp-fe-02.html; Table 2. Characteristics of COVID-19 Cases with reported on-set dates of 6 – 17 Feb 2020 (n=163).

##### Note from the authors of this paper

There are 115 passenger cases. 23 of them were in cabins *with* a confirmed case; and 52-92 of them were in cabins *without* a confirmed case. There was uncertainty in the data, hence a range was given, but in calculating the total, the largest number was used (23+92 = 115). Additional confirmed cases were also added to the 23 cases in cabins with a confirmed case, but the onset date was unknown, hence not considered in our calculation.

#### Onset data 3

Nishiura (2020) also presented a dataset. The paper was received by the journalon Fen 25, 2020, hence it supposed to be the most updated as compared to the official NIID or MHLW data.

##### Summary

There are 150 data. The three data sets differ significantly. Prof Nishiura data has the largest number of close contact category, much larger (103) than the other two; while the other two data sets had only a very small number of close contact cases.

The three sets of data are summarized in Figure S1.

**Figure S1.**
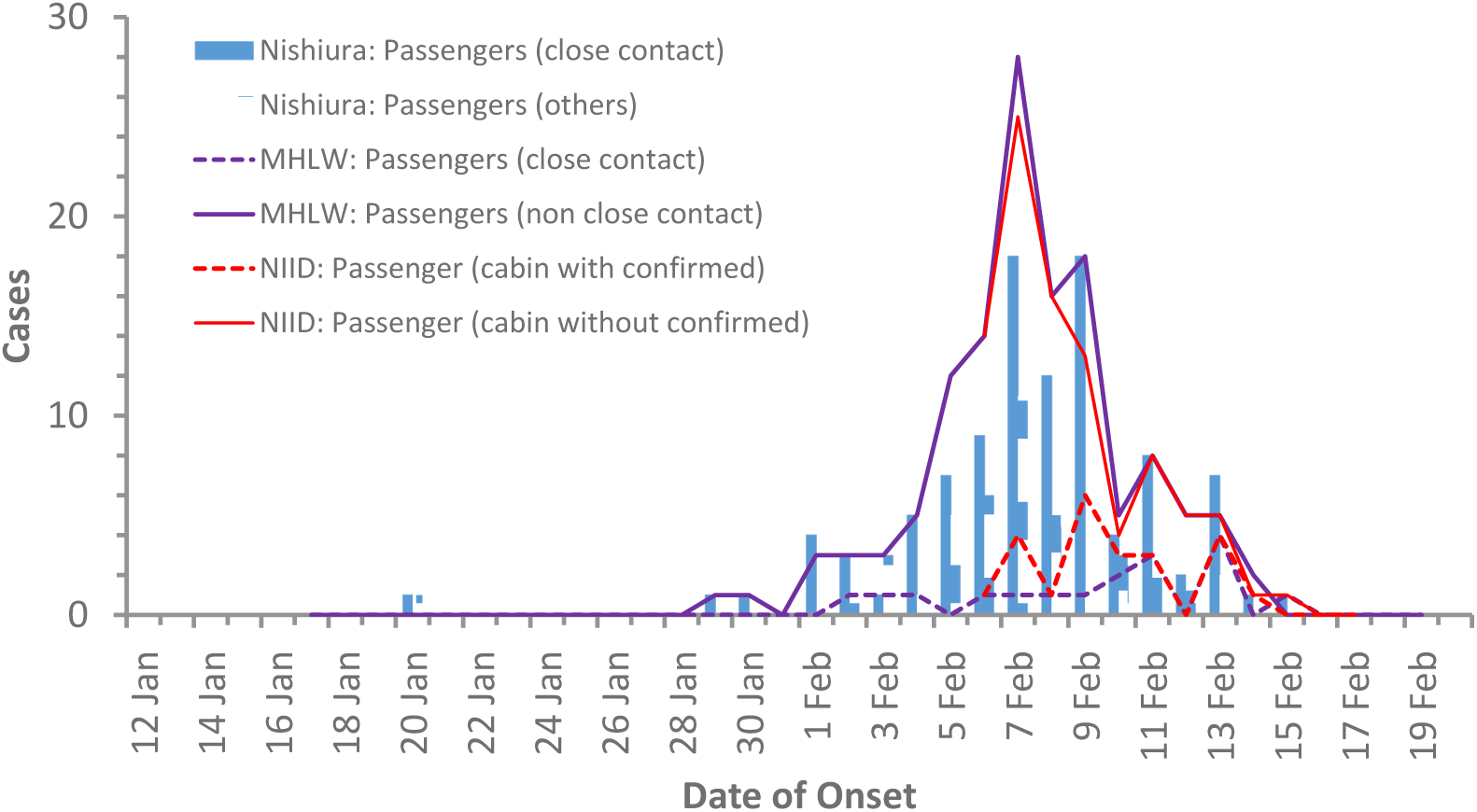
A summary of the three data sets of daily onset of symptoms among two categories of passengers. Close contact (those who stayed in a cabin room with an infected) and non-close contact or others (those who stayed in a cabin room without an infected)

“Non-close contact” and “Passengers in cabins without a confirmed case” in the MHLW and NIID datasets referred to as “the first confirmed case in each cabin”. The “passengers with close contact” in the dataset of Prof Nishiura seems to refer to “passengers in the infected cabins” (Takashita, personal communication). This explains that the number of close contact by Prof Nishiura was larger than MHLW and NIID if the second case was confirmed in each cabin.

We chose to use the official data in the main text in this study.

The predicted infection using the data set of Nishiura (2020) is shown in **Figure S2**. The predicted infection trend for crew is nearly identical with that shown in **Figure 1A**, with a peak on Feb 7. Unfortunately, we did not detect any difference between the “close contact” and “others” passengers, except that the “close contact” passengers did have infection on Feb 6 in Figure S2, i.e. after the quarantine starting date.

**Figure S2.**
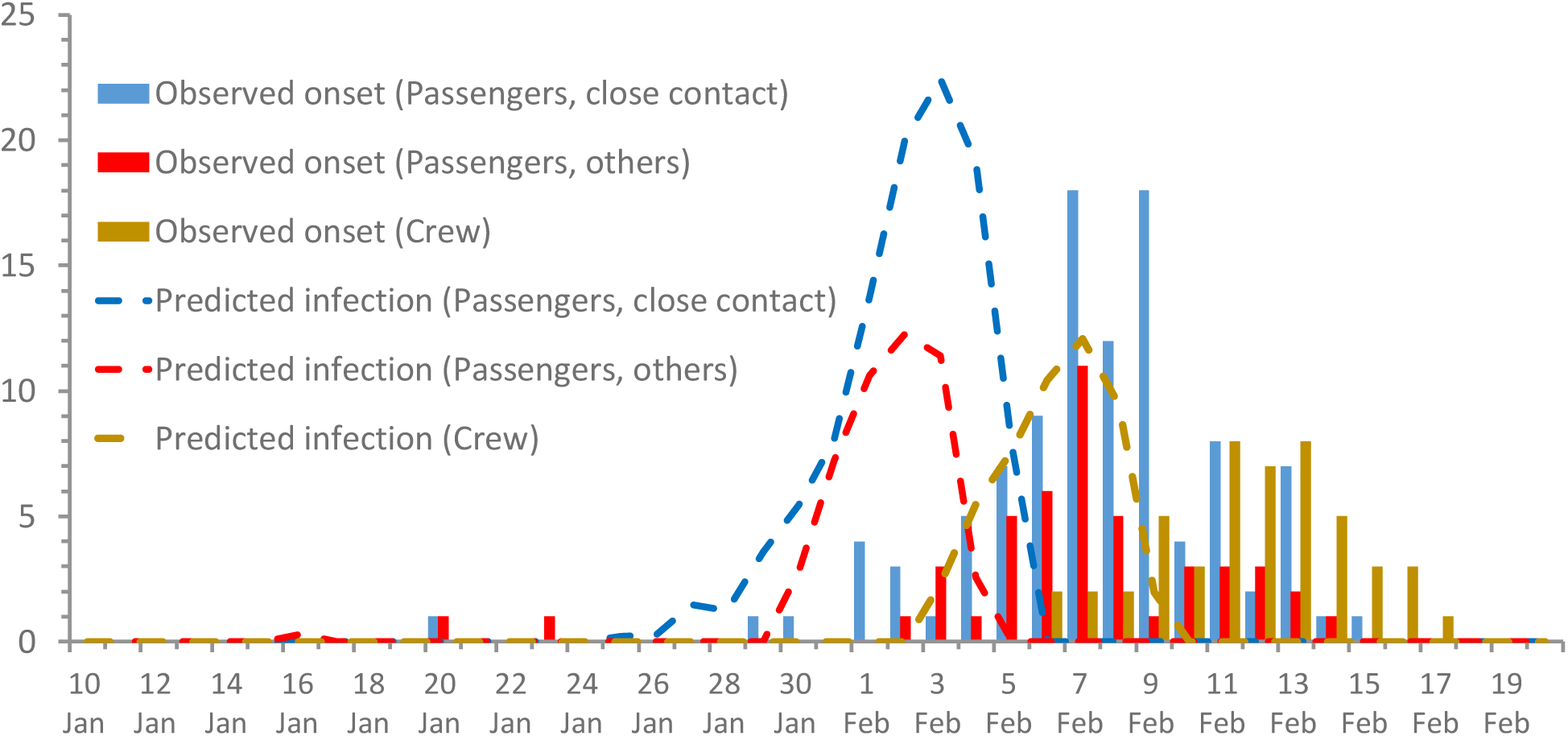
Predicted infection for the passengers (in two categories, close contact and others. Close contact refers in staying together with an infected individual in the same room), and crew.

